# Psychosocial vulnerabilities in newly arrived asylum seekers to the Greek island of Lesvos: a retrospective cross-sectional analysis

**DOI:** 10.1101/2023.11.01.23297929

**Authors:** Gregory Kavarnos, Miriam Bressaglia, Sally Hargreaves, Marie Norredam, Apostolos Veizis

## Abstract

**Background:** Kara Tepe Registration and Identification Camp on Lesvos Island in Greece is a controlled access facility with people in various stages of the asylum procedure housed in tents, rub halls and containers in poor living conditions for prolonged periods of time. However, data are lacking on psychosocial vulnerabilities and experiences of these populations, particularly the growing number of single women residing in these settings.We aimed to explore psychosocial needs among this incamped population.

**Methods:** We did a retrospective cross-sectional chart review of sociodemographic and psychosocial vulnerabilities of all migrants (>18 years of age) who were detained in Kara Tepe between 1st February 2021 and 31st January 2023. Data were extracted from the Knack Online Database used by INTERSOS HELLAS.

**Results:** 701 individuals were referred to the clinic for psychosocial work-up over a 2 year follow-up period, of which 165 subsequently received psychological support after initial health assessments. The majority (92.0%) of the cohort were women who were single, most from Afghanistan, Somalia, Congo, and Syria, with a range of medical and social vulnerabilities. 19.8% of the population had been subjected to gender-based violence including in their country of origin, during the migration route to Lesvos, or while on the island of Lesvos. The most common GBV incident reported was rape (48.9%) followed by physical assault (23.7%) and non-penetrative sexual assault (21.5%). 7% of the population presented with suicidal, or self-harming, behaviour, of which 26.2% had attempted suicide.

**Conclusion:** This study highlights for the first time the stark range of vulnerabilities experienced by predominantly single women in camp settings in Greece.

## Introduction

Since the implementation of the EU-Turkey deal in 2016 [1], the tens of thousands of asylum seekers arriving on Greek Islands from the coast of Turkey have largely been contained on these islands until their asylum claims are processed, with Lesvos continuing to be the epicentre of sea crossings from Turkey to Greece [2]. On the island of Lesvos, the majority of arrivals are currently placed in the main Registration and Identification (RIC) Camp of Kara Tepe, after the Moria camp burnt down in September 2020, leaving 12,000 people stranded on the streets of the island. Kara Tepe camp is a controlled access facility with people in various stages of the asylum procedure housed in tents, rub halls and containers in poor living conditions for prolonged periods of time [3]; with non-government organisations (NGOs) and charities involved in activities ranging from the provision of daily food supplies [4] and clothing, to medical and psychosocial support. Reports have highlighted a range of mental health issues in the migrant population of Lesbos, including self-harm and suicidal behaviour linked to past traumatic experiences, disruptions to their life, uncertainty, and loss of social and family networks, alongside significant barriers to accessing appropriate mental health care and treatment [3,5,6]. During the COVID-19 pandemic these camps experienced higher rates of COVID-19 than the host population, and longer periods of lockdown [7].

However, data are lacking on the specific vulnerabilities and experiences of these populations, particularly the growing number of single women residing in these settings [8]. The INTERSOS HELLAS Mental Health and Psycho-Social Service (MHPSS) was set up in February 2021, with the aim of offering psychosocial support to refugees and asylum seekers on the island of Lesvos. A team of psychologists, case workers, and cultural mediators worked outside of the RIC, and accepted referrals to the service from a large network of actors supporting the provision of housing, social and medical services.

## Methods

We did a retrospective cross-sectional chart review of socio-demographic and psychosocial vulnerabilities of all migrants (>18 years of age) who were: i) detained in the Kara Tepe Registration and Identification Camp in Lesvos Island and ii) referred to the INTERSOS HELLAS MHPSS programme between 1^st^ February 2021 and 31^st^ January 2023. A cultural mediator and/or an interpreter assisted with the interviews when needed. Data were extracted from the Knack Online Database used by INTERSOS HELLAS. The data came from two main sources: i) the initial assessment/intake interview performed at INTERSOS’ facilities by the social worker, and ii) the assessment and follow-up sessions of the psychologist. While every individual attending the service had an assessment by the social worker, not every patient went on to receive ongoing case management or psychological support.

We assess sociodemographic data including: age, sex, marital status and country of origin as well as self-reported psychosocial vulnerabilities including genderbased violence and mental health conditions. Descriptive data analysis was conducted using Looker Studio (formerly Google Data Studio). The study is reported according to STROBE guidelines [9].

As part of the initial assessment process, individuals were given a full explanation of the assessment interview, consent process and asked if their data could be used for reporting including research purposes. Oral and written consent was obtained. All data was fully anonymised before the research study took place, with individuals only identifiable by a randomly generated ID variable.

## Results

701 individuals were referred to the clinic for psychosocial work-up over a two-year follow-up period, of which 165 subsequently received psychological support after initial health assessments. The majority (92%) of the cohort were women and most were single (74.0%). The mean age was 30.31 years (range 13-75 years), and 48 (6.8%) were aged >50 years. Multiple countries of origin were identified, with most coming from Afghanistan (41.5%) followed by Somalia (21.5%), Congo (20.8%), Syria (3.8%), with other nationalities represented including Sierra Leone, Cameroon, Eritrea, Sudan, Cote d’Ivoire, and Angola. Individuals reported being in different stages of the asylum procedure: i) waiting for an interview to start asylum case (28.0%); ii) asylum case pending (24.5%); iii) rejected in the 1st and 2^nd^ instance (24.2%); iv) granted refugee status (5.6%); and v) other (17.7%) which includes: being a recipient of subsidiary status, or being in the process of making an appeal (e.g. awaiting further paperwork to prove their case, trying to find the funds to make a subsequent appeal).

19.8% of the population had been subjected to gender-based violence (GBV) of whom the majority were women (91.3%). They reported that gender-based violence had taken place in various settings, including in their country of origin, during the migration route to Lesvos, or while on the island of Lesvos, and 15% had experienced more than one assault. Furthermore, 6% of the total 701 people presented with acute mental health symptoms at the first assessment, such as hyperarousal, audial and visual hallucinations, disorientation, and mood irregularities. The majority of those with acute mental health symptoms (76.2%) presented clusters of symptoms which are not associated with any particular psychiatric disorder. Lastly, 7% of the population presented with suicidal, or self-harming, behavior. Out of these (26.2%) had attempted suicide.

Table 1 highlights the range of psychosocial vulnerabilities reported. The most common GBV incident reported was rape (48.9%) followed by physical assault (23.7%) and non-penetrative sexual assault (21.5%).

**Table 1.**
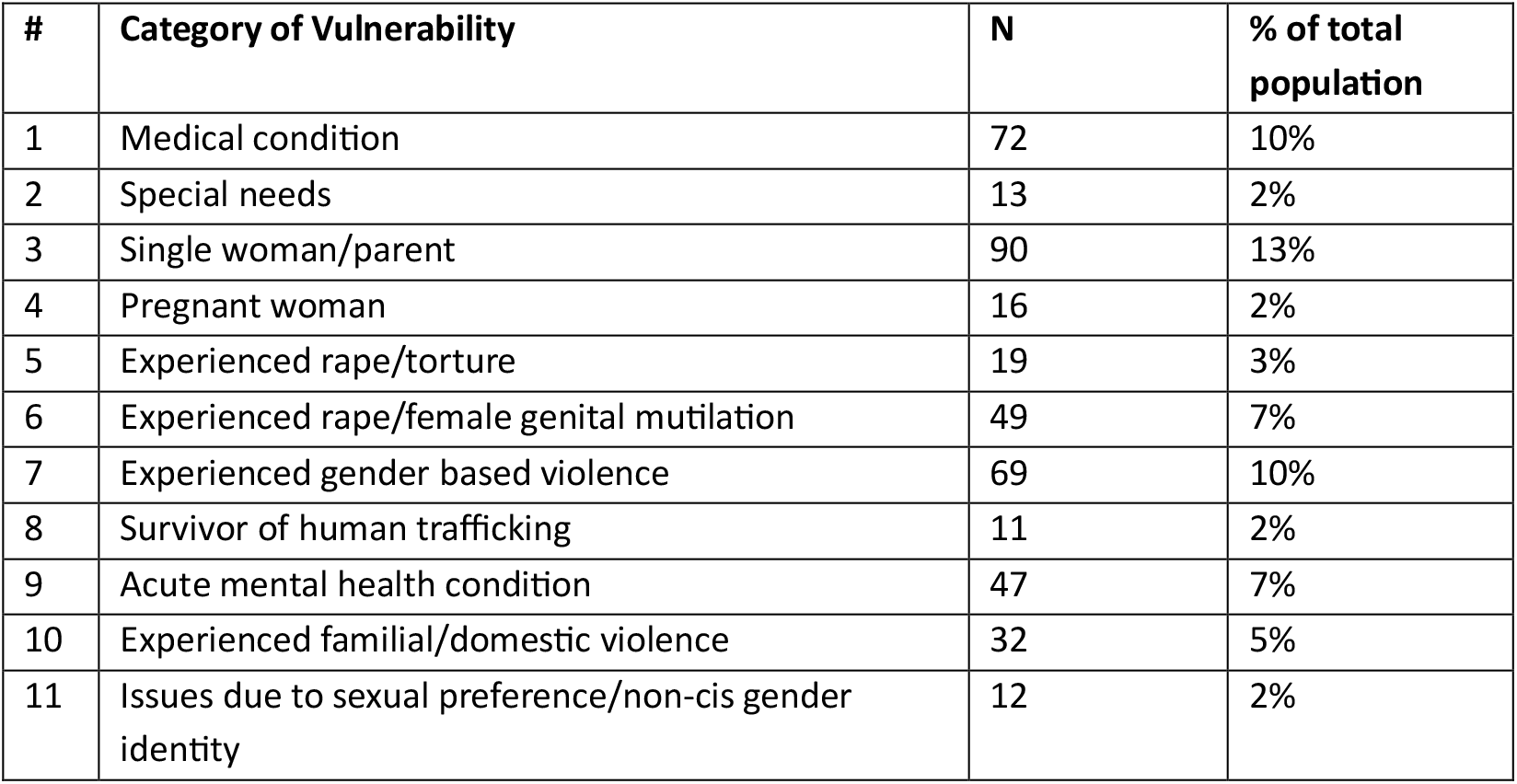
Category of vulnerabilities in the population. The n value in the table is the number of vulnerabilities reported, not the number of individuals reporting the incidents. As such, an individual may report more than one vulnerability. Categories #1-8 are recognized by the Greek Asylum Service as vulnerabilities which affect the asylum process.

## Discussion

This study highlights for the first time the stark range of vulnerabilities experienced by predominantly single women in camp settings, on the Greek islands. Levels of gender-based violence and suicidal ideation were high in our cohort. As well as their past histories, and the impact of the migration journey, these individuals also experience psychosocial distress and social suffering because of their uncertain and disrupted lives and the loss of social and family networks over many months in detention-like settings.

Previous studies have highlighted a range of mental health issues negatively associated with length of stay in camps, predominantly among men who in the past formed the majority of migrants in these settings. In one study from the Moria camp among 634 refugees (average age 23.2 years, predominantly Syrian and Afghan males) acute mental health crises were significantly and independently associated with the length of stay in the camp (p = 0·011; average length of stay in camp 70.9 days) Other studies too have confirmed a deterioration in mental health in Lesvos linked to containment in camps settings [3,12].

Concurrent with the containment issues, there is also a lack of long-term mental health services to deal with the severe trauma experienced by refugees and asylum seeker. There also needs to be a serious reassessment (and application) of the vulnerability criteria in the asylum process, as many people, although they fulfil vulnerability criteria [13], receive rejections for their asylum claims. Nonbinary and/or non-cis gender identification and the impact that these have on the life of the individual are not even part of vulnerability criteria, even when there are cases of life-threatening discrimination in an asylum seekers country of origin (https://www.humandignitytrust.org/lgbt-the-law/map-of-criminalisation/). In many cases GBV experiences, or the fact that the asylum seeker is a single woman, are not considered at all when vulnerability is being assessed.

As such the Ministry of Migration and Greek Asylum Service need to reassess the range of vulnerabilities being assessed, and direct Asylum Service staff to determine the presence of vulnerabilities more adequately during asylum interviews. This is necessary to make informed asylum application decisions, thus reducing the need for time consuming appeals during which the applicants are essentially prisoners.

## Contributors

AV and GK designed the study and ran the psychosocial programme. GK extracted and analysed the data with support from SH, MN and AV. GK wrote a first draft of the paper and all authors were involved in commenting on subsequent drafts.

## Methods

All methods were carried out in accordance with relevant guidelines and regulations. The protocol was approved by a INTERSOS medical unit and the ethical board

## Informed consent

Informed consent was obtained from all subjects.

## Data Availability

All data produced in the present study are available upon reasonable request to the authors

